# Can self-testing be enhanced to hasten safe return of healthcare workers in pandemics? Random order, open label trial using two manufacturers’ SARS-CoV-2 lateral flow devices concurrently

**DOI:** 10.1101/2024.04.04.24305332

**Authors:** Xingna Zhang, Christopher P Cheyne, Christopher Jones, Michael Humann, Gary Leeming, Claire Smith, David M Hughes, Girvan Burnside, Susanna Dodd, Rebekah Penrice-Randal, Xiaofeng Dong, Malcolm G Semple, Tim Neal, Sarah Tunkel, Tom Fowler, Lance Turtle, Marta Garcia-Finana, Iain E Buchan

## Abstract

**Background:** Covid-19 healthcare worker testing, isolation and quarantine policies had to balance risks to patients from the virus and from staff absence. The emergence of the Omicron variant led to dangerous levels of key-worker absence globally.

We evaluated whether using two manufacturers’ lateral flow tests (LFTs) concurrently improved SARS-CoV-2 Omicron detection and was acceptable to hospital staff. In a nested study, to understand risks of return to work after a 5-day isolation/quarantine period, we examined virus culture 5-7 days after positive test or significant exposure.

**Methods:** Fully-vaccinated Liverpool (UK) University Hospitals staff participated (February-May 2022) in a random-order, open-label trial testing whether dual LFTs improved SARS-CoV2 detection, and whether dual swabbing was acceptable to users. Participants used nose-throat swab Innova and nose-only swab Orient Gene LFTs in daily randomised order for 10 days. A user-experience questionnaire was administered on exit. Selected participants gave swabs for viral culture on Days 5-7. Cultures were considered positive if cytopathic effect was apparent or SARs-COV2 N gene sub-genomic RNA was detected.

**Results:** 226 individuals reported 1466 pairs of LFT results. Tests disagreed in 127 cases (8.7%). Orient Gene was more likely (78 cf. 49, P=0.03) to be positive. Orient Gene positive Innova negative result-pairs became more frequent over time (P<0.001). If Innova was swabbed second, it was less likely to agree with a positive Orient Gene result (P=0.005); swabbing first with Innova made no significant difference (P=0.85).

Of 311 individuals completing the exit questionnaire, 90.7% reported dual swabbing was easy, 57.1% said it was no barrier to their daily routine and 65.6% preferred dual testing. Respondents had more confidence in dual c.f. single test results (P<0.001).

Viral cultures from Days 5-7 were positive for 6/31 (19.4%, 7.5%-37.5%) and indeterminate for 11/31 (35.5%, 19.2%-54.6%) LFT-positive participants, indicating they were likely still infectious.

**Conclusions:** Dual brand testing increased LFT detection of SARS-CoV-2 antigen by a small but meaningful margin and was acceptable to hospital workers. Viral cultures demonstrated that policies recommending safe return to work ∼5 days after Omicron infection/exposure were flawed. Key-workers should be prepared for dynamic self-testing protocols in future pandemics.

**Trial registration:** https://www.isrctn.com/ISRCTN47058442 (IRAS:311842)

## Background

The Covid-19 pandemic stretched health systems worldwide.[1,2] Healthcare workers suffered high rates of infection and mortality,[3–5] and policymakers faced dilemmas in balancing risks. In late 2021, as Omicron hit the UK, hospitalised patients faced potentially greater risks from care-staff shortages (Figure 1) than from Covid-19.[6–10] Omicron’s increased transmissibility and immune evasion demanded a rethink of Covid-19 policies for healthcare workers and the public.[10–13]

**Figure 1.**
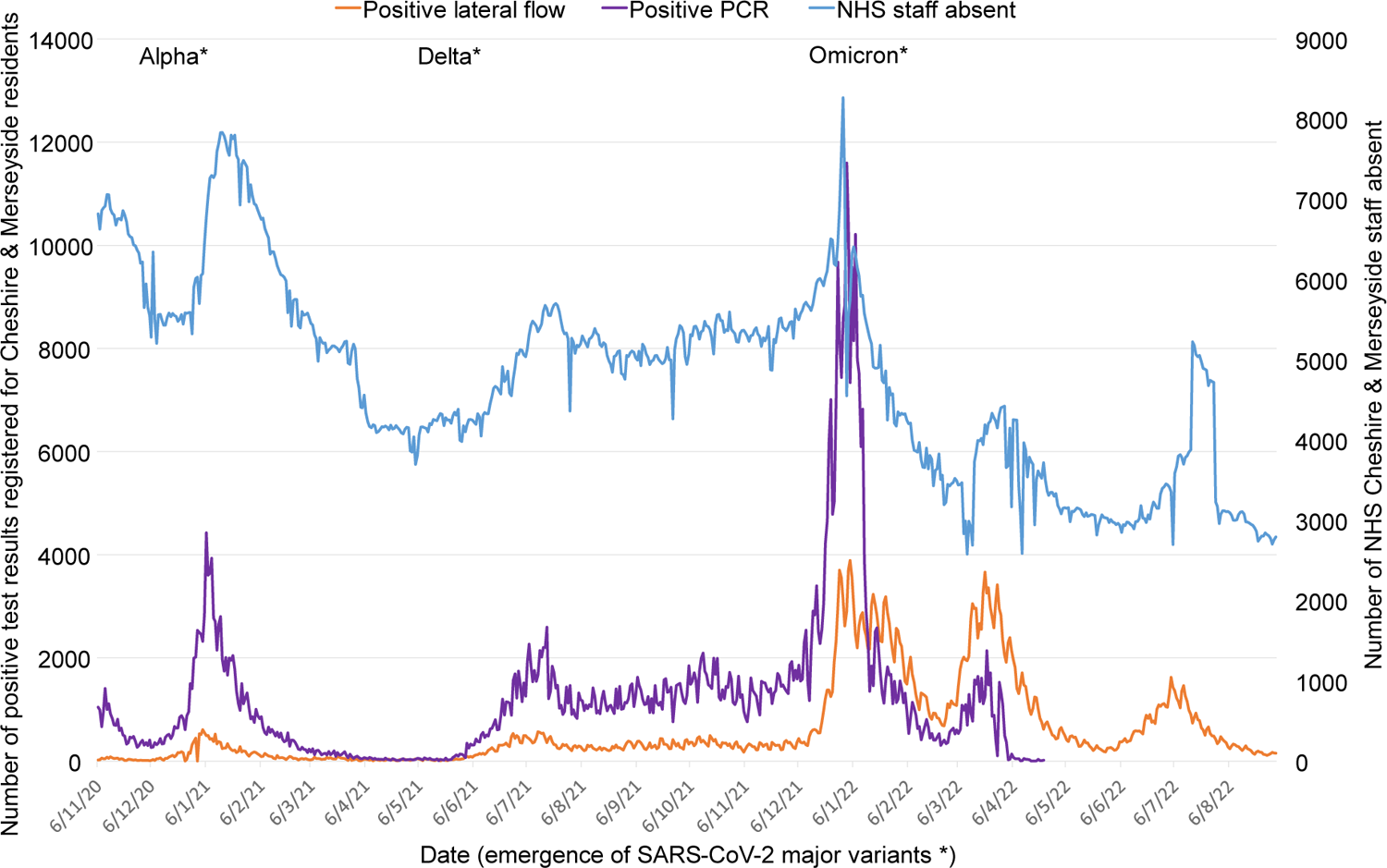
Numbers of NHS staff absent, and numbers of positive PCR and lateral flow test results reported for residents of Cheshire & Merseyside, UK from the start of introduction of lateral flow community testing to the end of the study period

Pre-Omicron, UK healthcare workers required a negative PCR 10 days from exposure to return from quarantine.[14,15] Waiting (typically 48-hours) for PCR results delayed return work,[15] and PCR capacity affected care-service continuity.[16,17] By December 2021, it was evident that SARS-CoV-2 lateral flow tests (LFTs) were reasonable and affordable indicators of infectiousness. LFTs from some manufacturers used nose-only swabbing, others nose-throat swabbing, with nose-only testing assumed to have better compliance. Policymakers were concerned that nose-only swabbing might delay detection of Omicron, which was reportedly shed from the throat ahead of the nose[18] – a concern not addressed by national testing quality assurance programmes.[19,20]

In December 2021 and January 2022, NHS staff testing policies changed to address staff shortages. Based on mathematical modelling, NHS workers were permitted to return from isolation or quarantine: after two consecutive days of negative LFTs beyond 5 days since exposure or first positive test; or if still testing positive, 10 days from symptom onset or first positive test, provided they felt well enough.[14,15,21] This guidance was updated on 7^th^ January 2022 to advise local risk assessments for those testing positive on days 10-14.[22]

The modelling of serial negative LFT results to inform return to work was performed by the Scientific Pandemic Influenza Group on Modelling (SPI-M)[23] and UK Health Security Agency (UKHSA)[24] alongside unpublished viral culture studies for the New and Emerging Respiratory Virus Threats Advisory Group (NERVTAG).

This study was commissioned by the UK Covid-19 Testing Initiatives Evaluation Board (TIEB) to extend its testing quality assurance programme. We investigated whether SARS-CoV-2 antigen detection in daily self-testing was improved by using kits from two manufacturers concurrently; one requiring nose-only and one nose-throat swabbing.[19] Real-world testing sensitivity and NHS staff acceptability were the main outcomes. A nested virus culture study assessed the infectiousness of individuals still testing positive after day-5 since symptom onset or first positive test, as the US policy was to return to work after day-5 without testing. Data from this study informed UK policies via TIEB.[25]

## Methods

### Aim

We aimed to evaluate effectiveness and acceptability of dual vs single brand SARS-CoV-2 antigen lateral flow self-testing among hospital workers, and to determine whether culturable SARS-CoV-2 Omicron was present 5-7 days after a positive test or significant exposure.

### Trial design

An open-label, randomised-order trial of using two LFT brands concurrently in daily self-testing with the ‘Test-to-release’,[26] or Daily Contact Testing design.[27–30]

### Setting

Participants comprised fully vaccinated NHS workers using Covid-19 staff-testing facilities for contacts or cases at Liverpool University Hospitals NHS Foundation Trust, UK. Participants entered the study via three routes (Appendix 1): i) test-negative but close contact; ii) test-positive asymptomatic; or iii) test-positive symptomatic. Staff booked a swab on-line where they received study information and consented to participate. Data were collected via on-line questionnaires and NHS record linkage.

### Intervention

The study used two LFT brands widely available via NHS Test & Trace in February 2022: the nose-only swab Orient Gene and nose-throat swab Innova (Xiamen Biotime Biotechnology) kits. These have similar performance curves vs viral load when compared to PCR results.[20]

Participants were asked to take two LFTs daily for 10 days, and on day-1 and day-5 to return swabs for quantitative PCR. Test order was detailed on an information sheet (Appendix 2), with daily LFTs in random order (Innova or Orient Gene first) and PCR on day-1 and day-5. Participants uploaded LFT results via NHS Test & Trace systems – enhanced with automated image reading for accuracy and ease of reporting.[31]

A nested study considered culture of viable virus at Days 5-7 from first positive test.

### Outcomes

The primary outcome was the discordance of results from concurrent LFTs. Secondary outcomes were participant compliance, and self-reported experience of dual c.f. single testing.

### Sample size

Calculations (see Appendix 3) assumed 18% drop-out and 10% test-positivity. The proportion of consented individuals not returning data was higher than expected (Figure 2), and test-positivity was >10%. Power to detect a difference between dual and single testing was the main target and the number of participants testing positive (n=167) was similar to the number required (n=164). It was later reported that SARS-CoV-2 LFTs were more sensitive to Omicron than prior variants, with Orient Gene more sensitive than Innova.[20]

**Figure 2.**
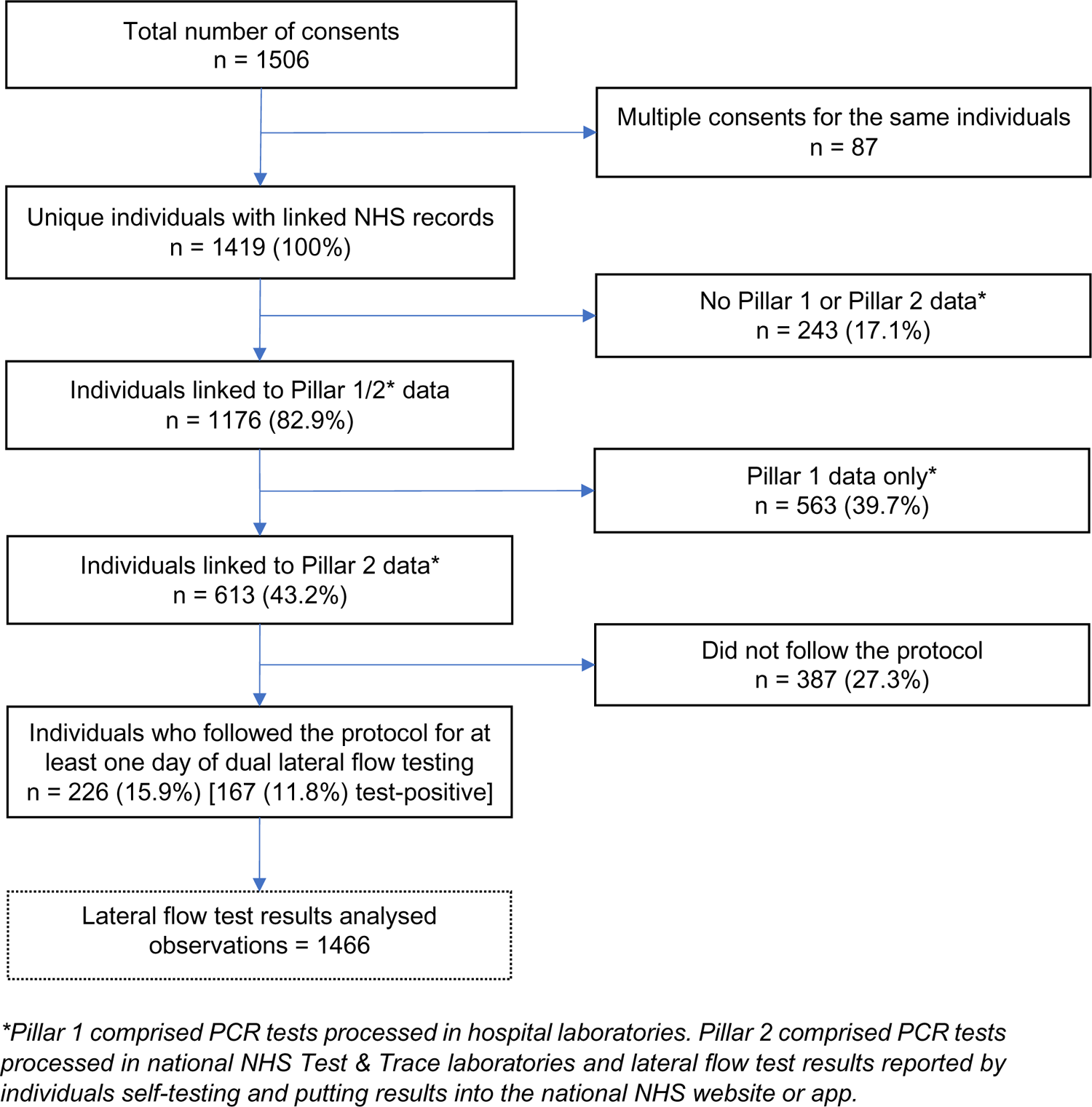
Flow of participants from consent to data analysed

### Viral culture and sequencing to determine lineage

Appendix 4 details viral culture, RNA-extraction, sequencing and bioinformatics methods used to infer the presence of replicable SARS-CoV-2 lineages from swab samples. In brief, Calu3 cells, cultured at 10^5 cells/well in 24 well plates, were inoculated for viral culture, incubated and checked for cytopathic effect (CPE) after three days. If CPE was visible supernatants were sampled for RNA extraction. If no CPE was visible a 2^nd^ passage was performed before supernatant sampling. RNA was extracted from supernatants and used for amplicon sequencing by MinION, using a published method. Fastq reads were analysed using the ARTIC[32] bioinformatic pipeline and lineages were called with Pangolin[33]. LeTRS was used to assess the presence of N gene sub genomic RNA (sgRNA)[34], indicative of active viral transcription.

### Statistical methods

Discordance of result-pairs from two LFT brands was analysed with McNemar’s test, including Yang’s adjustment and logistic mixed-effects models to account for test-clustering within individuals over time and in study-day groups.[35] Trends over time in discordance were analysed with a logistic mixed-effects model addressing clustering within individuals with study-day groups disaggregated.

Comparison of users’ confidence in single vs dual testing from questionnaire ordinal score data used a Wilcoxon signed ranks test and exact confidence interval as score distributions were skewed. Confidence intervals for binomial proportions used the Clopper-Pearson method, and for logistic mixed-effects models the Wald method. Analyses were performed using R version 4.3.1. Results were verified independently by two statisticians. Results are presented as main effect with 95% confidence interval.

### Patient and public involvement

UK HSA’s Research Ethics Governance of Public Health Practice Group (EGG), including lay members, along with TIEB, fed back on drafts of the study protocol as part of the approvals process. Additional public involvement over data governance was provided by Liverpool City Region Civic Data Cooperative.

## Results

### Main outcomes

Data were collected between 7^th^ February and 8^th^ May 2022. 226 participants reported at least one day of dual LFT results between study day-1 and day-10, giving 1466 pairs of tests, of which 127 (8.7%) were discordant. Figure 2 shows the flow of participants from consent to analysis, Figure 3 the recruitment patterns over time.

**Figure 3.**
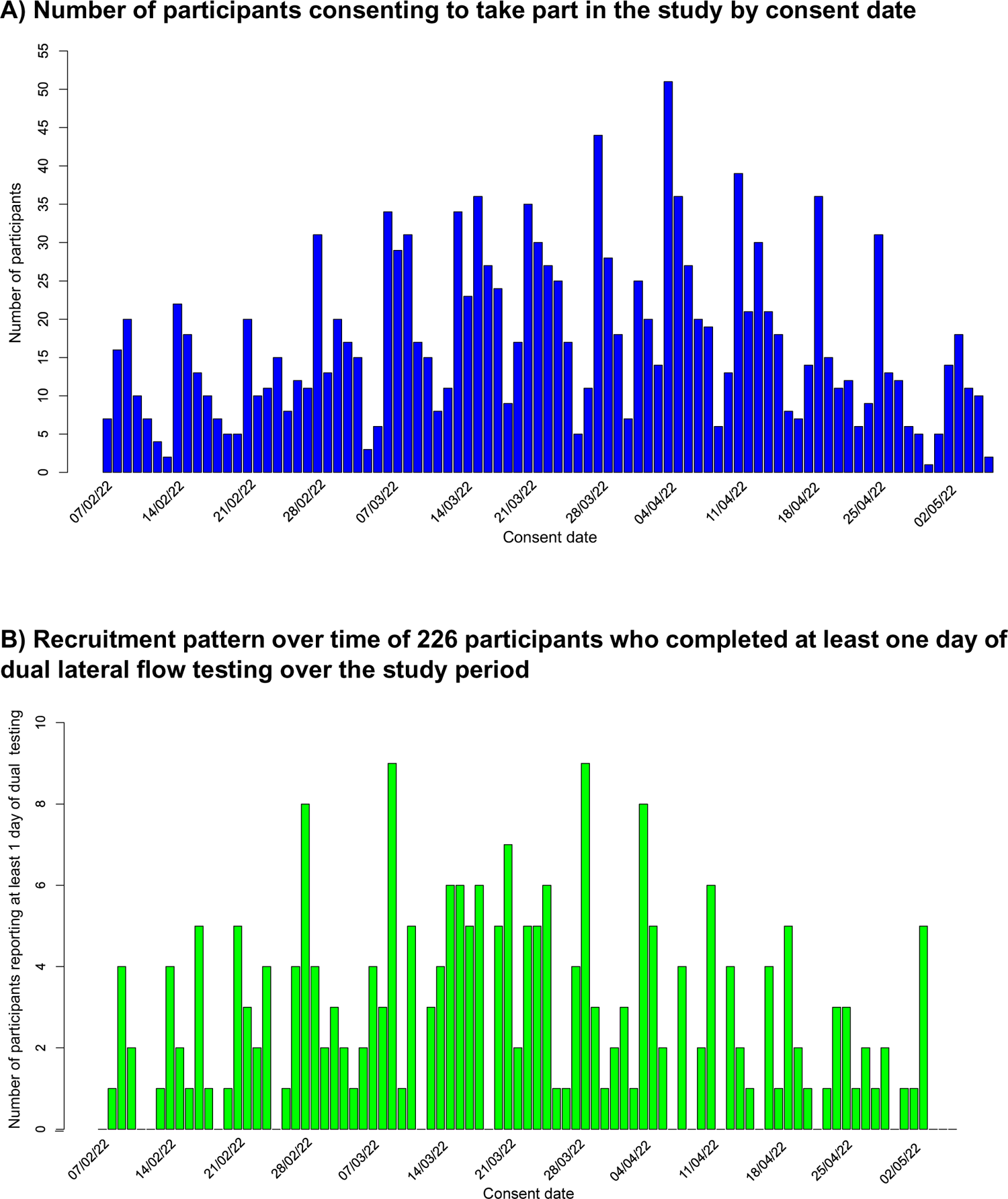
Overall, Orient Gene had double the odds of being positive compared to Innova when the two tests disagreed (Table 1). 156 (93.4%; 88.5%-96.7%) suspected infections were detected with Orient Gene compared to 163 (97.6%; 94.0%-99.3%) with Innova. Out of the test-positive cohort of 167, 59 (35.3%) had at least one subsequent period of 2 or more consecutive days of dual negative LFTs. Of these, none had a pair of positive LFT results afterwards.

**Table 1.** Lateral flow test results by brand.

|  |  | Innova |  | TOTAL |
| --- | --- | --- | --- | --- |
|  |  | Negative | Positive |  |
| Orient Gene | Negative | 596<br>(40.7%) | 49<br>(3.3%) | 645 |
|  | Positive | 78<br>(5.3%) | 743<br>(50.7%) | 821 |
| TOTAL |  | 674 | 792 | 1466<br>(100%) |

*Test pairs with any equivocal result were excluded. McNemar (Yang-adjusted) $\chi^2=4.636$ , $P=0.03$ , logistic mixed-effects model for discordant tests odds ratio (OR)=2.1(1.1-4.1), $P=0.03$ .*

When Orient Gene was the first test (Table 2), Orient Gene positive Innova negative was a more likely discordant result than Innova positive Orient Gene negative (OR=2.7, 1.3-5.2; P=0.005). No significant difference was observed when Innova was the first test (OR=1.1, 0.5-2.3; P=0.85, Table 3). Direct comparison of discordant test pairs shows the odds of an Orient Gene positive with Innova negative discordance was 4.5 times higher when Orient Gene was first vs Innova first (OR=4.5, 1.1-18.1; P=0.04).

**Table 2.**
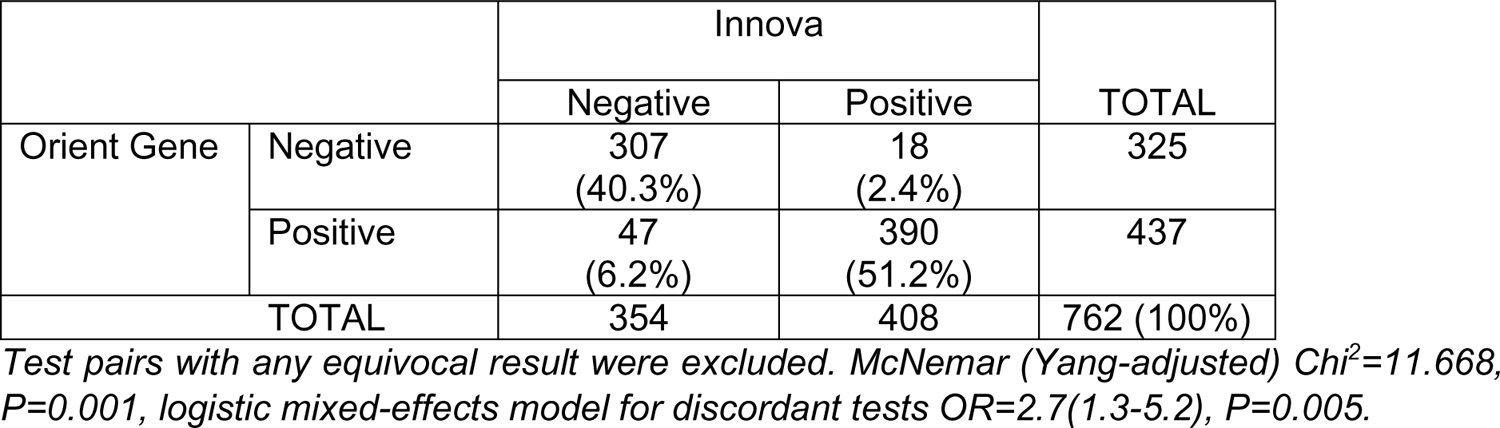
Lateral flow test results when Orient Gene was recorded first.

|  |  | Innova |  | TOTAL |
| --- | --- | --- | --- | --- |
|  |  | Negative | Positive |  |
| Orient Gene | Negative | 307<br>(40.3%) | 18<br>(2.4%) | 325 |
|  | Positive | 47<br>(6.2%) | 390<br>(51.2%) | 437 |
| TOTAL |  | 354 | 408 | 762 (100%) |

*Test pairs with any equivocal result were excluded. McNemar (Yang-adjusted) $\chi^2=11.668$ , $P=0.001$ , logistic mixed-effects model for discordant tests OR=2.7(1.3-5.2), $P=0.005$ .*

**Table 3.** Lateral flow test results when Innova was recorded first.

|  |  | Innova |  | TOTAL |
| --- | --- | --- | --- | --- |
|  |  | Negative | Positive |  |
| Orient Gene | Negative | 289<br>(41.1%) | 31<br>(4.4%) | 320 |
|  | Positive | 31<br>(4.4%) | 353<br>(50.1%) | 384 |
| TOTAL |  | 320 | 384 | 704 (100%) |

*Test pairs with any equivocal result were excluded. McNemar (Yang-adjusted) $\chi^2 < 0.001$ , $P > 0.99$ , logistic mixed-effects model for discordant tests OR=1.1(0.5-2.3), $P=0.85$ .*

Of the167 participants who tested PCR or LFT positive at study entry or became LFT test-positive during the study (Table 4), the proportion of Orient Gene positive discordant tests increased significantly over time (OR: 1.2, 1.1-1.3; P<0.001), and not significantly for Innova (OR: 1.1, 0.97-1.2; P=0.15). Direct comparison of the two discordant groups using a logistic mixed effects model did not show statistical significance (OR: 1.2, 0.99-1.6; P=0.07), however, small numbers of discordant groups (Figure 4) may have limited the power to resolve this effect.

**Figure 4.**
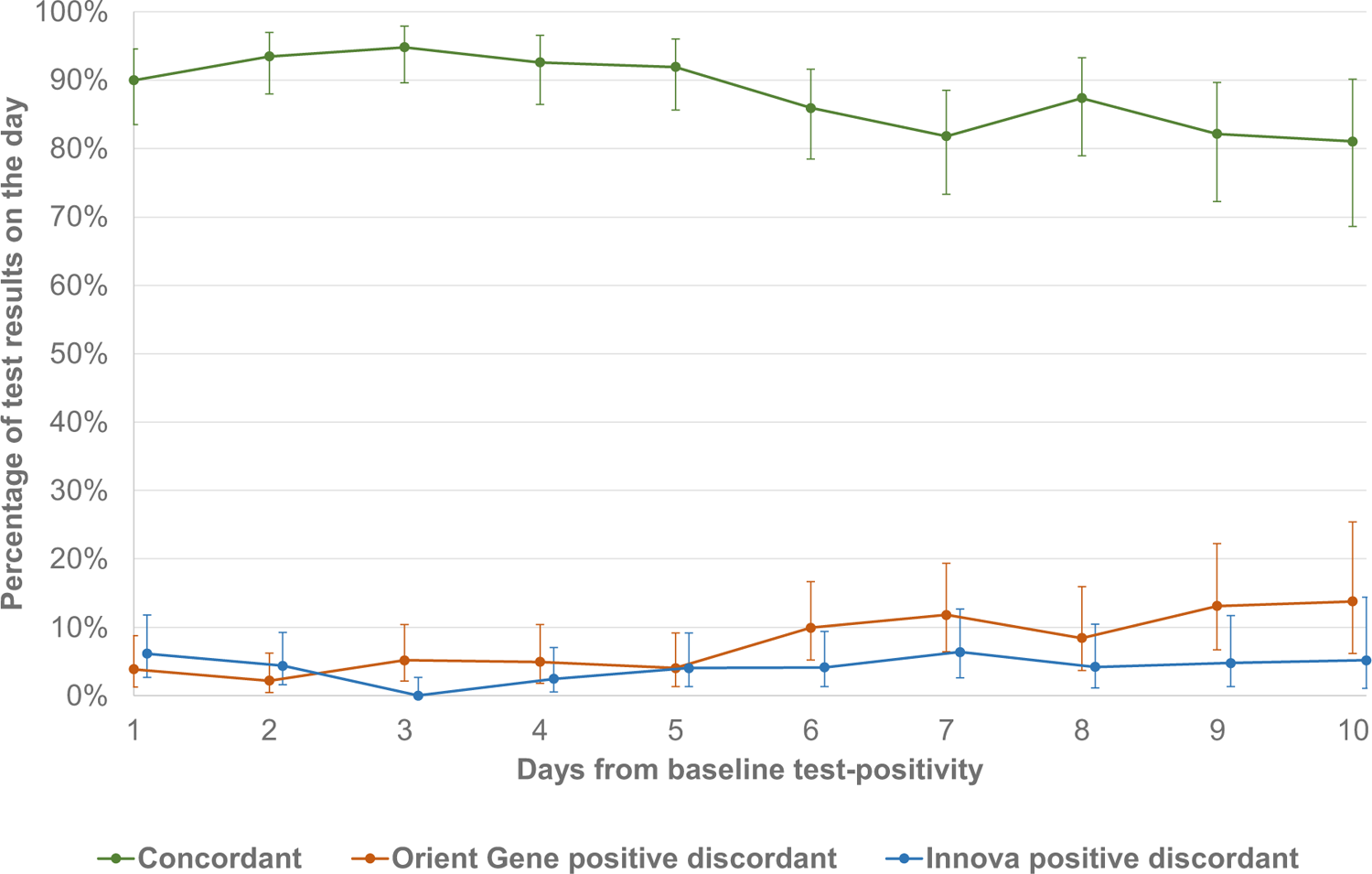
Percentage of concordant and discordant lateral flow test result pairs by brand and days from baseline (study entry or first positive test) for those testing positive

**Table 4:**
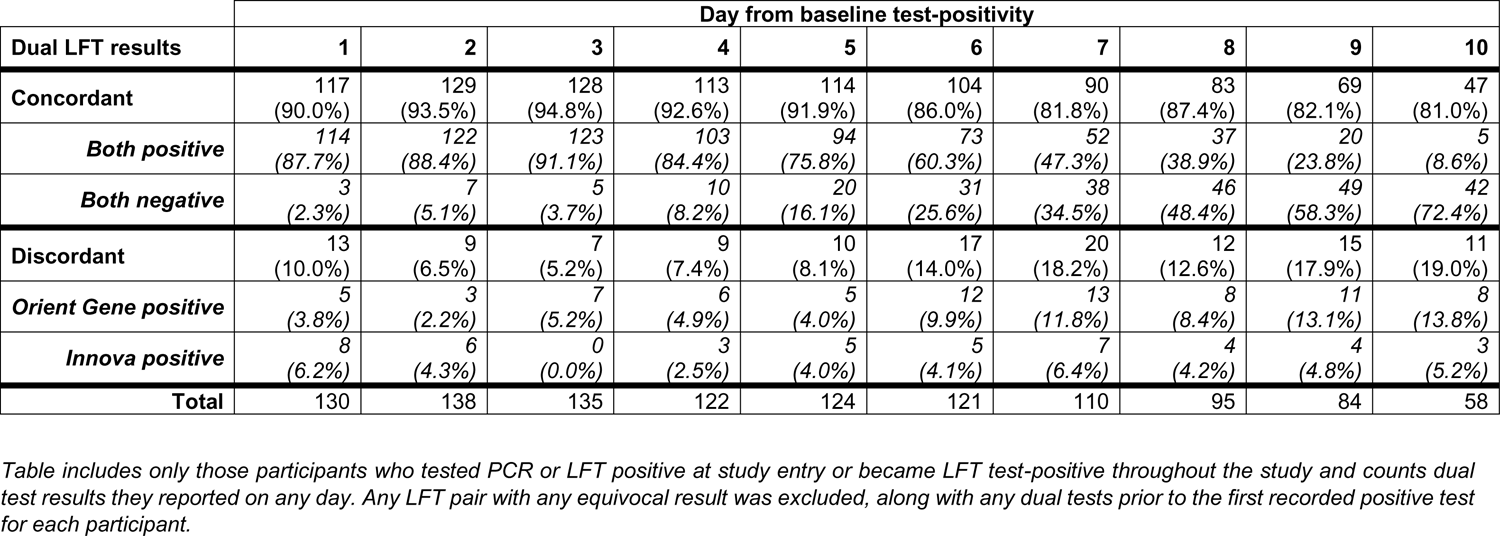
Dual lateral flow test (LFT) results by brand and days from baseline (study entry or first positive test) for individuals who tested positive.

A total of 125 individuals had a positive PCR test at recruitment/consent. In this group, the first reported LFT result pairs were both positive for 116 (92.8%), both negative for 3 (2.4%), Innova positive only for 3 (2.4%) and Orient Gene positive only for 3 (2.4%) – a discrepancy rate (∼detection uplift from dual vs single testing) of 6/125 (4.8%, 1.8%-10.2%). For the second day of available results, 99 were positive on both LFTs (90%), 5 were negative on both (4.5%), 5 were Orient Gene positive only (4.5%), and 1 was Innova positive only (1%) – a discrepancy rate of 6/110 (5.5%, 2.0%-11.5%). Considering two days of consecutive test results, only 2 (1.6%) individuals would have been LFT negative and PCR positive within 48 hours.

### Viral culture analysis

Viral cultures were analysed for 41 participants (see Appendix 4 for details): 31 continuing LFT positive and 10 reverting negative on days 5-7. 6/31 (19.4%, 7.5%-37.5%) of the continued LFT positives were culture positive, 9/31 (29.0%, 14.2%-48.1%) were indeterminate by cytopathic effect (CPE), none of these had N-sgRNA detected by sequencing. Two additional cultures with no CPE had N-sgRNA detected, both at low level. 2/10 (20.0%, 2.5%-55.6%) of the reverting LFT negatives were indeterminate by CPE with N-sgRNA not detected by sequencing. Two cultures without CPE had N-sgRNA detected at low levels. PCR was carried out on the original swab aliquot: all came back SARS-CoV-2 positive, with S Gene target present in 36 (likely Omicron BA.2) and absent in 5 (likely Omicron BA.1).

### Exit questionnaire survey

311 participants responded to the exit survey between 10^th^ February and 20^th^ July 2022. 229 (73.7%) identified as a woman, 77 (24.7%) as a man, 0 as non-binary and 5 (1.58%) preferred not to say. Professions were: 57 (18.5%) doctors; 84 (27.2%) nurses; 78 (25.0%) allied health professionals; 25 (8.2%) clinical support staff; 44 (14.1%) administration/clerical staff; 22 (7.07%) other staff. When asked “how easy was the swabbing process” 154 (49.6%) selected “very easy”, 128(41.2%) “easy”, 26(8.4%) “neither easy nor difficult”, 3(0.9%) “difficult”, and none “very difficult”. When asked “How much of a barrier is having to take a throat as well as a nose swab for your daily rapid test?” 178(57.1%) selected “not at all”, 80 (25.6%) selected “slight barrier”, 38(12.3%) selected “somewhat of a barrier”, 12 (3.94%) selected “moderate barrier”, and 3 (1.0%) selected “extreme barrier”. When asked “Would you fit taking two rapid tests into your daily routine within an hour of leaving for work?” 90 (28.9%) respondents selected “definitely”, 58(18.6%) selected “very probably”, 110 (35.3%) selected “prob ably”, 44(14.22%) selected “probably not”, and 9 (2.9%) selected “definitely not”. When asked “If you were asked to continue to do two tests daily instead of one, would you?” 204 (65.6%) selected “yes”, 57(18.2%) selected “no”, and 50(16.2%) selected “no reference”. When asked “which mode of testing are you most confident about” and given a rating scale from 1= “no confidence” to 10= “full confidence” for “single test” and “double test” the median scores were 8 for single test and 9 for double test – a median difference of 1 (0.5 to 1; P<0.001).

### Discussion Main findings

We found that practical combination of LFT brands with different swab types enhanced antigen detection differently at different time points during infection. Combining Orient Gene and Innova LFTs improved SARS-CoV-2 (Omicron BA.1/2) antigen detection by a small, but meaningful, amount compared to a single test.

Orient Gene was more likely to be the sole positive test – with Orient Gene positive Innova negative results becoming more frequent over successive days. If Innova was swabbed second it was less likely to agree with a positive Orient Gene result; swabbing first with Innova made no significant difference.

Dual testing was largely acceptable to hospital staff, with most reporting dual swabbing was easy. Almost two-thirds preferred to continue dual testing if required and had more confidence in dual cf. single testing results. Over half said dual swabbing was no barrier to daily routines.

Almost a fifth of individuals with positive LFTs at days 5-7 had positive culture, and a further third had indeterminate culture, indicating they were likely still infectious.

### Comparison with other studies

Evidence on end-to-end risk mitigation using LFTs in the Covid-19 pandemic is limited, with a few small serial testing and viral culture studies nested within studies comparing LFT and PCR performance.[20,36,37] Ours is the only real-world trial, to our knowledge, of combining LFTs to enhance risk-mitigation in balancing risks from Covid-19 vs healthcare staff shortages. The shorter incubation period of Omicron compared with earlier variants challenged previous risk-mitigations[38,39] and there were concerns over later nasal shedding invalidating nose-only swab LFTs,[18] which we showed were mitigated by dual testing.

Viral culture studies with early variants found that people infected with SARS-CoV-2 became infectious 1-2 days before the onset of symptoms and remained infectious until 7 days later.[38,40] Our data showed a substantial proportion of individuals were potentially infectious beyond this point. Most work on this topic has placed too much emphasis on the median duration of infection at the expense of considering variability and its impact on fixed time-period policies for return from isolation or quarantine.

### Strengths and limitations of this study

This is the first study, to our knowledge, to compare dual with single brand LFT self-testing of healthcare workers in managing the risks from Covid-19 vs under-staffed care due to high numbers isolating or quarantined. It was performed when the UK was under pressure from Omicron variants in early 2022. It was thus a realistic test of enhanced risk-mitigation and comprehensively considered LFT sensitivity, participant experience, and security (using viral cultures) of prompt return to work. The 10-day observation period allowed assessment of the contemporary Covid-19 policies for healthcare worker release from isolation and quarantine. The results give insights into the combined performance of two brands of LFT, which cannot be inferred from the usual comparison with concurrent PCR tests.[20,41]

Our study had limitations: slow research approvals meant the peak of the initial Omicron epidemic was missed. Staff burnout and low morale slowed recruitment and prolonged the study duration. National policies for NHS staff testing changed several times during the study,[13,42] and public access to LFTs was reduced on 1^st^ April 2022,[42] potentially also impeding participant uptake. These barriers reflect the real-world challenges of evaluating new risk-mitigations two years into a pandemic and under winter pressures.

Only 16% of consented participants followed the protocol for at least one day – this likely limits the representativeness of the survey results. Finally, viral culture interpretation was limited – parallel PCR swabbing and sequencing would have given more confidence that SARS-CoV-2 was present and not another virus.

### Policy implications

Throughout the Covid-19 pandemic, policies for testing healthcare workers changed many times.[10,13,16,43] In the UK, LFT self-testing became part of life. However, with the rise of Omicron policymakers feared that nose-only LFT swabbing may miss numerous infections. Our study allayed these fears, showing reasonable concordance of the widely available Orient Gene nose-only and Innova nose-throat swab LFTs. Policymakers were therefore right to use all available stocks of LFTs, including those with nose-only swabs, to mitigate elevated risks.

Despite pandemic pressures, study participants reported largely positive experiences of using two LFTs instead of one for daily self-testing. The improvement in detection from dual testing was small yet meaningful in a universal health system coordinating risk-mitigations system-wide.

Internationally, there was pressure to balance risks from Covid-19 with those from mass absence of key workers. Our viral culture study shows that fixed isolation time policies, such as the US advice to return to work 5 days after testing positive, were flawed. The UK’s ‘test-to-release from isolation’ (after two days of negative LFT results) policy, formed in December 2021, was reasonable given staffing pressures at the time.

The speed, convenience, and socialisation of LFT self-testing in the UK allowed enhanced Covid-19 risk-mitigation under pressure from Omicron. A better UK response would have extended testing quality assurance from public health agencies to the NHS. Ideally, more serial samples of daily antigen, nucleic acid and culturable virus testing would have informed policy modelling. Future pandemic preparations globally should consider closer surveillance of serial self-testing to inform evolving risk-mitigations.

## Conclusion

Policymakers’ fears that nose-only LFT swabbing may miss a substantial proportion of Omicron BA.1/2 infections were allayed by our study of NHS workers in the UK between February and June 2022. Combining the widely available Innova nose-throat and Orient Gene nose-only LFT kits increased Omicron detection and was acceptable to participating hospital staff self-testing in isolation or quarantine. This improvement was small yet meaningful in a universal health system coordinating risk-mitigations system-wide. The US policy of return to work 5 days after testing positive was shown by our viral culture results to be flawed. The speed, convenience, and public socialisation of LFT self-testing in the UK allowed enhanced Covid-19 risk-mitigation during the pressures caused by the Omicron variant. A better UK response would have extended testing quality assurance from public health agencies to the NHS. Future pandemic preparedness may be enhanced by continuous surveillance of serial self-testing, considering end-to-end risk mitigation as well as technology performance.

## Ethical approval

The study protocol was developed with the UK Covid-19 Testing Initiatives Evaluation Board (TIEB) and approved by UK Health Security Agency (UKHSA) Urgent Studies Ethics Committee. TIEB was part of the UK Covid-19 testing initiatives evaluation programme, which included academics and public health professionals independent of this study. The motivation for the study came from a request by Merseyside Resilience Forum to UKHSA and NHS England to vary Covid-19 testing policies in response to dangerous levels of NHS staff absence in December 2021. TIEB signed off the study protocol on 4^th^ January 2022 and UKHSA Research Support and Governance Office approved the study on 25^th^ January 2022 as NR0308. The sponsor code for this study is UoL001685 and trial registration code IRAS ID 311842; https://www.isrctn.com/ISRCTN47058442.

## Supporting information

Graphical abstract

## Data Availability

The study required person identifiable data and the main analyses were conducted on a de-identified extract. The fully anonymised data for reproducing the results are available from https://www.isrctn.com/ISRCTN47058442. The study protocol can be downloaded from https://github.com/iain-buchan/cipha/blob/master/SMART_Release_Return.pdf and statistical analysis plan from https://github.com/iain-buchan/cipha/blob/master/SMART_RR_SAP.pdf.

## Acknowledgements

We thank all participants and wider staff at Liverpool University Hospitals NHS Trust who supported the study, Cheshire & Merseyside Blood Bikes volunteers who made home-based viral culture swab delivery and collection possible, the UK Testing Initiatives Evaluation Board for their generous support with urgent preparation of the protocol and discussion over interim findings to support policymaking, and UKHSA Research Ethics and Governance Group. Thanks to Kathryn Scott for proofreading this manuscript.

## Contributors

IB, XZ and CC drafted the manuscript. IB drafted the protocol, led the study and the preparation of reports and this paper. CC drafted the statistical analysis plan and led data management with GL. CC, DH, GB, SD, MGF and IB ran and corroborated three independent statistical analyses. CS led the study management. TN ran the trial site. TF and ST ran the UKHSA operations for the trial. LT and CJ ran the viral culture sub-study. RPR and XD performed viral sequencing and bioinformatic analyses. Each author contributed substantial intellectual content during manuscript drafting or revision and accepts accountability for the overall work by ensuring that questions pertaining to the accuracy or integrity of any portion of the work are appropriately investigated and resolved. All authors approved the final version of the manuscript. IB is the study guarantor. The corresponding author (IB) affirms that all the listed authors meet the authorship criteria and that no others meeting the criteria have been omitted.

## Affirmation

IB affirms that the manuscript is an honest, accurate, and transparent account of the study being reported; that no important aspects of the study have been omitted; and that any discrepancies from the study as planned have been explained.

## Dissemination to participants and related patient and public communities

Findings were presented to the UK Testing Initiatives Evaluation Board and associated patient and public involvement groups throughout the study and in final form in September 2022. TN, as infection prevention and control lead and trial site (Liverpool University Hospitals) lead has disseminated results to participants.

## Provenance and peer review

The study was commissioned by the UK’s Department of Health and Social are. The work has been peer reviewed by external reviewers in the UK Testing Initiatives Evaluation Board at each stage, from protocol formation to presentation of findings.

## List of abbreviations

Covid-19 (coronavirus disease 2019); SARS-CoV-2 (severe acute respiratory syndrome coronavirus 2); LFT (lateral flow test); CPE (cytopathic effect); NHS (UK National Health Service); TIEB (UK Covid-19 Testing Initiatives Evaluation Board); UKHSA (UK Health Security Agency); NERVTAG (New and Emerging Respiratory Virus Threats Advisory Group); CDC (US Centres for Disease Control and Prevention).

## Appendix 1: Participant Pathways

The full study protocol is available from: https://github.com/iain-buchan/cipha/blob/master/SMART_Release_Return.pdf

### A. Uninfected contact participant pathway (illustrated in Figure A1.1)

1. Household member of NHS worker was notified they were Covid positive, so their NHS contact started quarantine and notified their employer.
2. Employer had adopted SMART Release & Return testing schedule as their local standard policy and directed the staff member to a booking website for the scheme, which provided information sheet, consent process and directions to the unit/site.
3. Participant received a 10-day pack of daily dual LFTs and 2 PCR home test kits, and if they had not had a positive Covid test in the past 90 days they took a swab for quick turnaround (binary) PCR.
4. Participant received PCR negative result on Day 0 and returned to work on Day 1 with DCT.
5. Either Innova (nose/throat) or Orient Gene (nose only) LFTs were taken each morning (or pre-shift) before breakfast in randomised order for 10 days – an information sheet in the pack directed the participant day by day. Either LFT reporting positive was an overall positive result.
6. On Day 1 the participant also took home a PCR swab (randomised order with the two LFTs) and returned it by post to Pillar 2 / other (ringfenced) Q-RT-PCR capacity, and the result was not used for any purpose other than research.
7. A second Q-RT-PCR swab was taken on Day 5.
8. Exit questionnaire gathered participant experiences.

**Figure A1.1.**
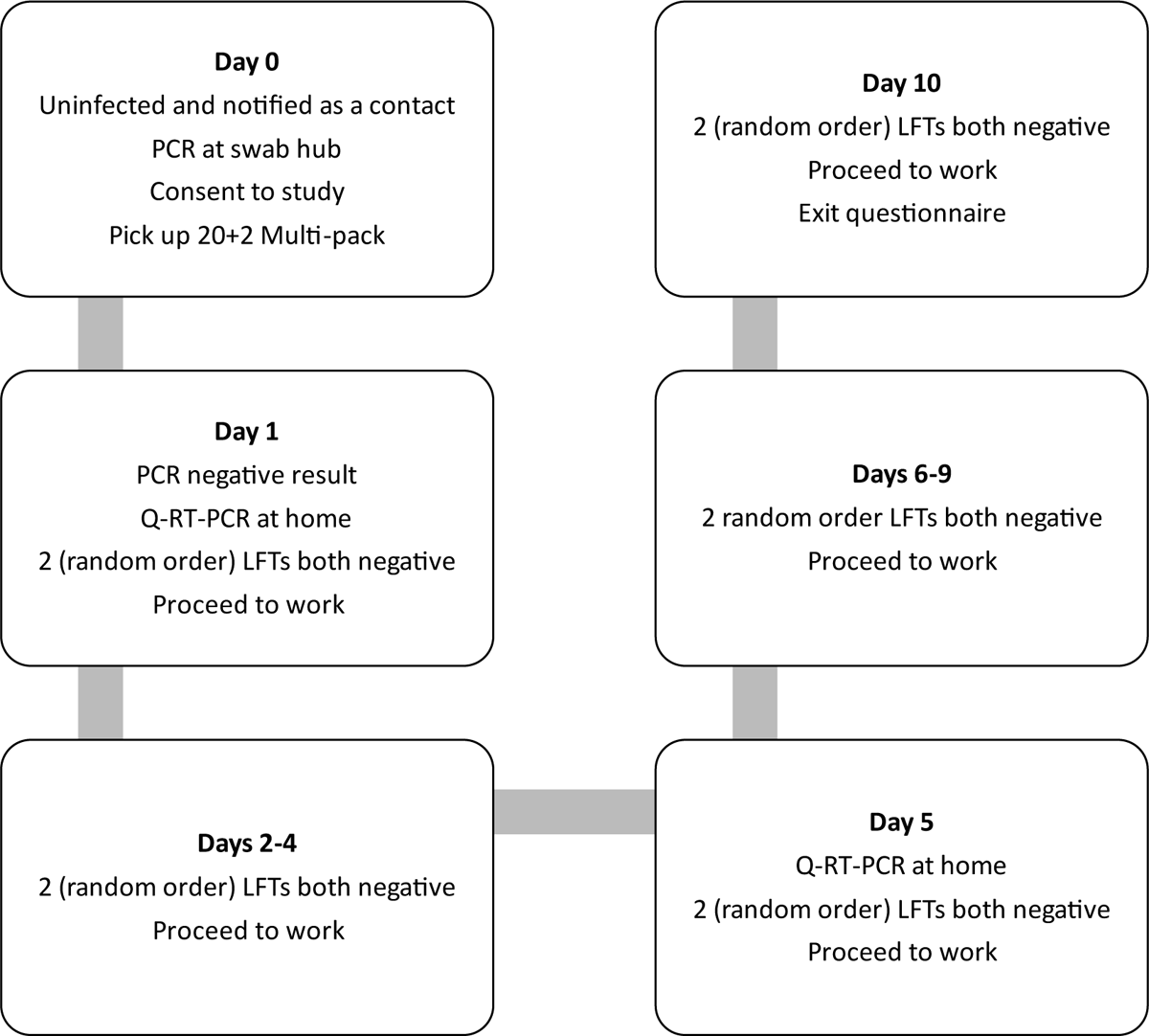
Workflow for participant who tested negative throughout the study

### B. Asymptomatic infected contact participant pathway (illustrated in Figure A1.2)

1. Household member of NHS worker was notified they were Covid positive, so their NHS contact started quarantine and notified their employer.
2. Employer had adopted SMART Release & Return testing schedule as their local standard policy, directed the staff member to a booking website for the scheme, and directed them to the standard testing/reception site.
3. Consented participant took quick turnaround (binary) PCR test to return to work from quarantine on DCT and received a 10-day pack of daily dual LFT + 2 PCR home test kits.
4. Participant received PCR positive result on Day 0 and stayed at home.
5. Either Innova (nose/throat) or Orient Gene (nose only) LFTs were taken each morning before breakfast in randomised order – an information sheet in the pack directed the participant day-by-day.
6. On Day 1 the participant also took home a PCR swab (randomised order with the two LFTs) and returned it by post to Pillar 2 / other (ringfenced) Q-RT-PCR capacity, and the result was not used for any purpose other than research.
7. Second Q-RT-PCR swab was taken on Day 5. (Participant was selected to be in the viral culture sample of 30 cases – and their swab in viral transport medium was collected from their home).
8. If Day 5 and 6 dual LFT results (4 tests) were negative the participant may return to work.
9. Daily dual LFT testing continued until Day 10.
10. If still testing LFT positive at Day 7 the participant was advised to call and arrange a RT-Q-PCR swab in viral transport medium for culture.
11. Exit questionnaire gathered participant experiences.

**Figure A1.2.**
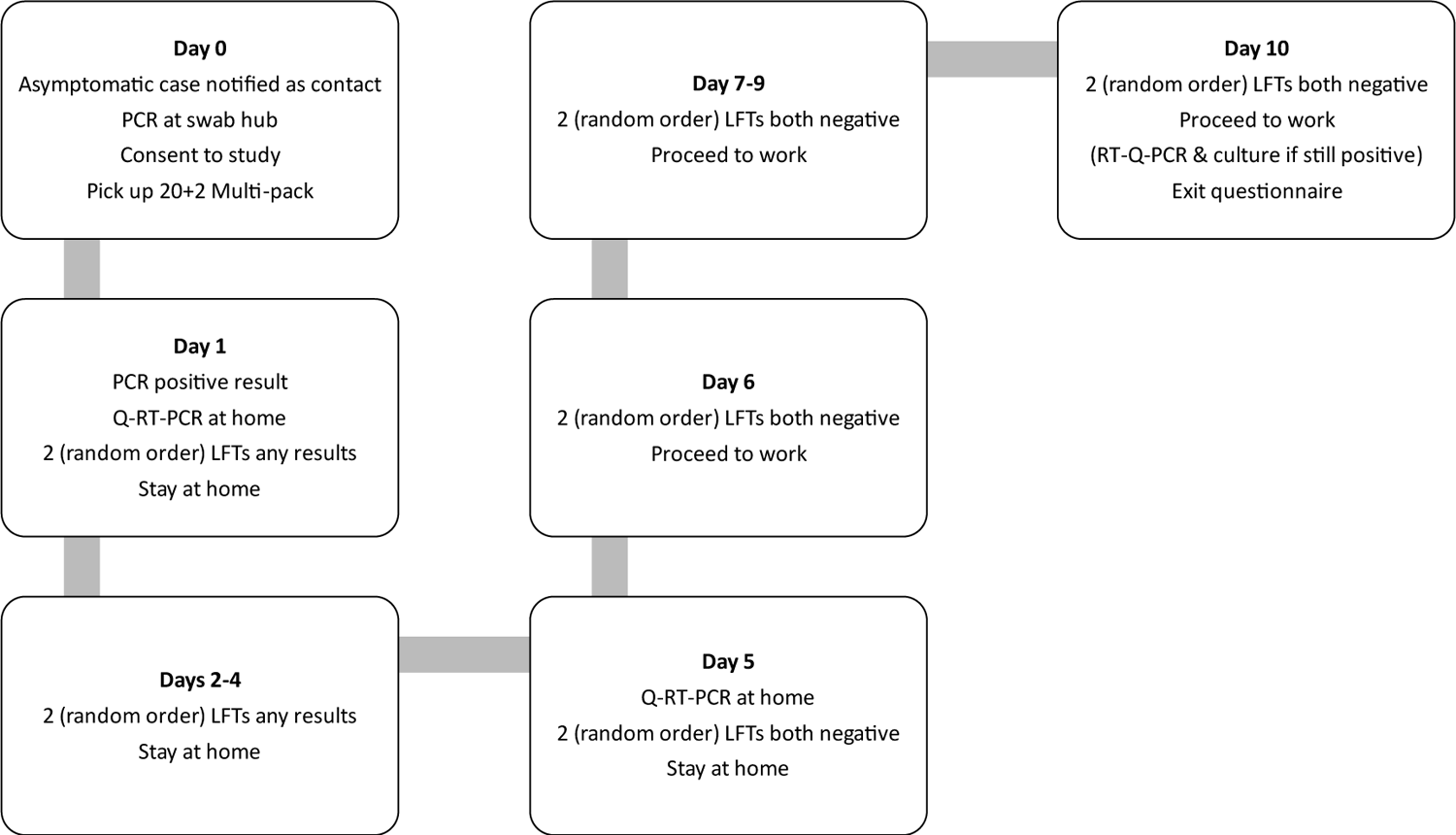
Workflow for contact who tested positive in the beginning of the study

### C. New case referred to the study (illustrated in Figure A1.3)

1. NHS worker was notified they were Covid positive and notified their employer.
2. Employer had adopted SMART Release & Return testing schedule as their local standard policy, directed the staff member to a booking website for the scheme, and directed them to the standard testing/reception site.
3. Consented participant received a 10-day pack of daily dual LFT + 2 PCR home test kits.
4. Innova (nose/throat) and Orient Gene (nose only) LFTs were taken each morning before breakfast in randomised order – an information sheet in the pack directed the participant day-by-day.
5. On Day 1 the participant also took home a PCR swab (randomised order with the two LFTs) and returned it by post to Pillar 2 / other (ringfenced) Q-RT-PCR capacity, and the result was not used for any purpose other than research.
6. Second Q-RT-PCR swab was taken on Day 5.
7. If Day 5 and 6 dual LFT results (4 tests) were negative the participant may return to work.
8. Daily dual LFT testing continued until Day 10.
9. If still testing LFT positive at Day 7 the participant was advised to call and arrange a RT-Q-PCR swab in viral transport medium for culture.
10. Exit questionnaire gathered participant experiences.

**Figure A1.3.**
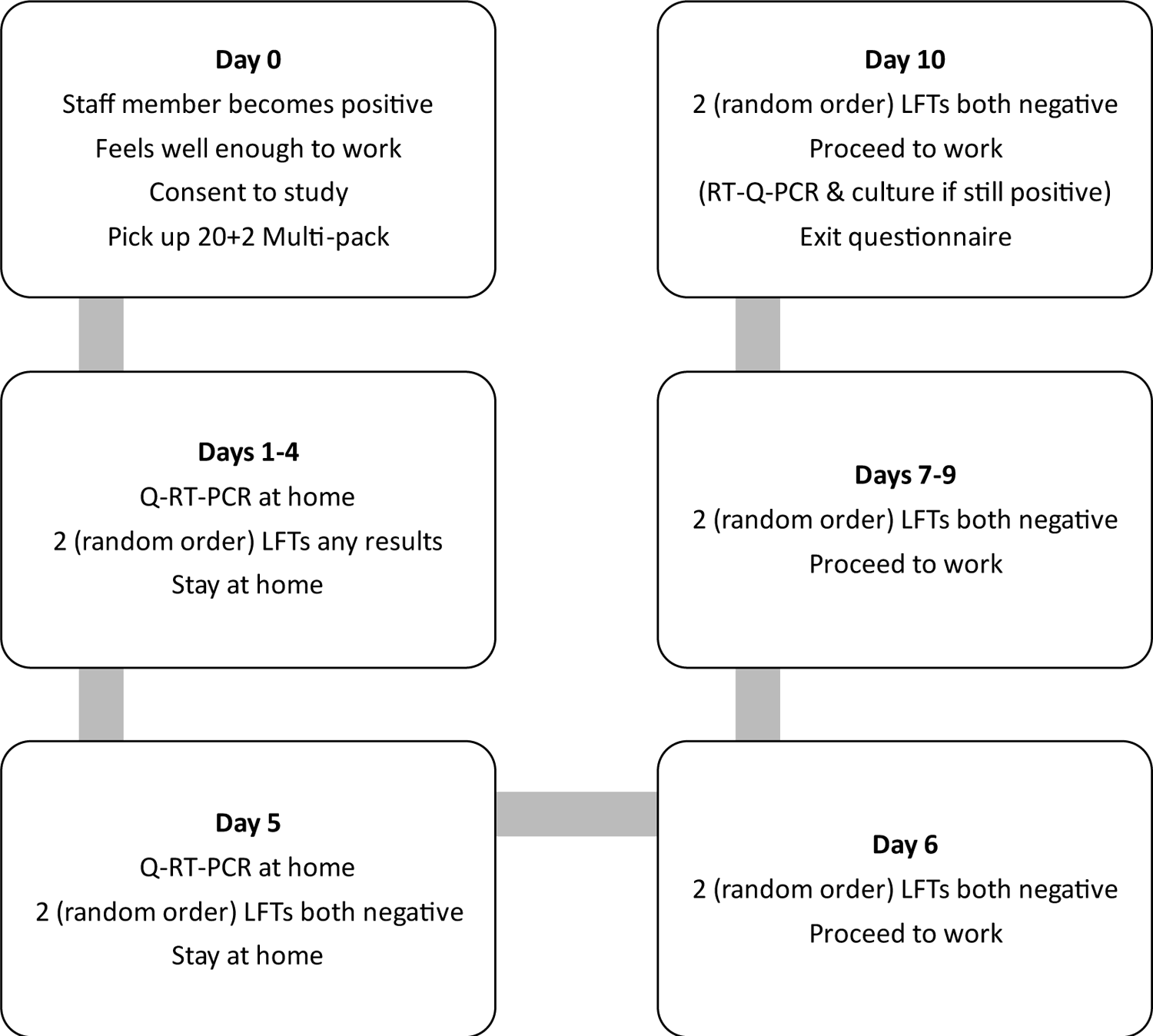
Workflow for participant who became positive at some point during the study or entered the study as a case

## Appendix 2: Participant information

A personalised randomisation sheet was inserted in the participants’ pack of swabs/tests with boxes/swabs marked A, B or C, with a typical schedule shown below…

The information booklet provided to all participants can be downloaded from: https://github.com/iain-buchan/cipha/blob/master/SMART_RR_Participant_Information.pdf

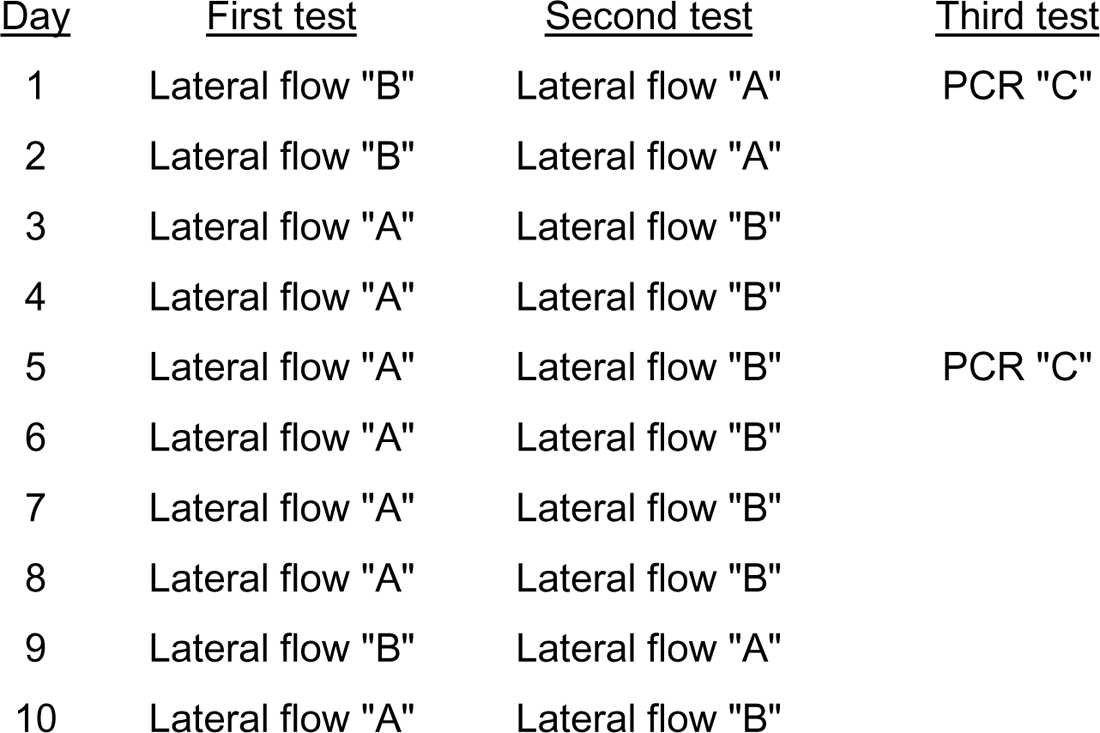

## Appendix 3: Sample Size Calculation

Assuming a conservative LFT sensitivity of 0.5 (accepting sensitivity varies as viral load changes over time) for PCR-LFT concordance, the proportion of cases missed on two consecutive days of single LFD testing would then be about 0.25 (0.5^2; with the limitation that within-individual physiological and behavioural factors may break independence). With dual testing this proportion over two days would be about 0.0625 (0.5^4), probably higher due to dependence between two consecutive LFTs done on the same day. We therefore assumed this proportion to be 40% higher (0.0625*1.4=0.09). So, assuming a LFT sensitivity of 0.5, for every 100 PCR positives, 25 cases are expected to be missed by two consecutive single tests versus approximately 9 cases with dual testing (i.e., 4 tests over two consecutive days).

The estimated odds ratio for single versus dual testing is then 3.37 ((0.25/0.75)/(0.09/0.91)) and the proportion of PCR positives with discordant pairs is about 16/100 positive cases (since the 9 negative cases by dual testing would be also negative by single testing).

Assuming these values for the underlying odds ratio and the % of discordant pairs between the two approaches, the number of positive cases required with 80% power at 5% significance to detect a significant difference between the two approaches in the proportion of cases missed is about 164 positive cases.[A3i] We raised it to 200 to account for loss to follow-up. Using only a contact cohort with about 10% case rate this would require a sample of 2000. We eventually recruited 1929 participants.

Table A3.1 shows the sample size for various values of sensitivity and for different scenarios regarding the dependence between two consecutive LFTs done on the same day. Note that the sample size required varies substantially with the within-individual correlation, which is unknown. We assumed some level of dependence between two tests conducted between minutes of each other, but not tests conducted on consecutive days, and had ignored the manufacturer factor.

Table A3.1 assumes that the two LFTs achieve the same level of sensitivity. Deviation from this assumption (i.e., allowing some degree of differentiation in sensitivities) is not expected to alter the sample size reported in Table A3.1 significantly. The power that can be achieved to detect a 15% drop in sensitivity over a two-day period with nose-only swabbing, when compared to nose-throat swabbing with a kit from a different manufacturer but with equivalent device sensitivity, is provided in Table A3.2 for different sample sizes and values of sensitivity. Subsequent reports of Innova and Orent Gene LFT sensitivity relevant to PCR showed similar profiles across different levels of viral load and different variants, but with Orient Gene the more sensitive.[A3ii-iii] Both tables use equation (7.1) in Machin et al. (2011)[A3i] based on discordance between results.

[A3i] Machin D, Campbell MJ, Tan SB, Tan SH. *Sample Size Tables for Clinical Studies*. John Wiley & Sons; 2011.
[A3ii] Eyre DW, Futschik M, Tunkel S, Wei J, Cole-Hamilton J, Saquib R, et al. Performance of antigen lateral flow devices in the UK during the alpha, delta, and omicron waves of the SARS-CoV-2 pandemic: a diagnostic and observational study. *The Lancet Infectious Diseases* 2023;23(8):922–32. Doi: 10.1016/S1473-3099(23)00129-9.
[A3iii] Performance of lateral flow devices during the COVID-19 pandemic. GOV.UK. Available at https://www.gov.uk/government/publications/lateral-flow-device-performance-data/performance-of-lateral-flow-devices-during-the-covid-19-pandemic. Accessed May 3, 2023.

**Table A3.1.** PCR positive sample size vs LFT sensitivity to detect a significant difference between dual and single LFT testing in the proportion of discordant cases with 80% power and 5% significance level.

| LFD Sensitivity | $P_s \equiv P_{\text{single}} = \text{Prob (negative LFT based on a single test \vert positive PCR)}$ | $P_s \equiv P_{\text{dual}} = \text{Prob (dual negative LFTs based on 2 consecutive tests within minutes \vert positive PCR)}$ | $P_1 = \text{Prob (negative LFTs on 2 consecutive days \vert positive PCR)} = P_s^2$ | $P_2 = \text{Prob (dual negative LFTs on 2 consecutive days \vert positive PCR)} = P_d^2$ | Odds Ratio | Proportion of discordant pairs | Number of PCR positives required |
| --- | --- | --- | --- | --- | --- | --- | --- |
| Assuming independence ($P_{\text{dual}} = P_{\text{single}} * P_{\text{single}}$) | | | | | | | |
| 0.7 | 0.3 | 0.09 | 0.09 | 0.0081 | 12.1 | 8.2% | 131 |
| 0.6 | 0.4 | 0.16 | 0.16 | 0.0256 | 7.25 | 13.4% | 99 |
| 0.5 | 0.5 | 0.25 | 0.25 | 0.0625 | 5.0 | 18.8% | 92 |
| 0.4 | 0.6 | 0.36 | 0.36 | 0.1296 | 3.78 | 23.0% | 98 |
| Assuming some level of dependence between two tests conducted within minutes |  |  |  |  |  |  |  |
| $P_{\text{dual}} = P_{\text{single}} * P_{\text{single}} * 1.2$ | | | | | | | |
| 0.7 | 0.3 | 0.11 | 0.09 | 0.0117 | 8.07 | 7.8% | 159 |
| 0.6 | 0.4 | 0.192 | 0.16 | 0.0369 | 4.98 | 12.3% | 142 |
| 0.5 | 0.5 | 0.3 | 0.25 | 0.09 | 3.37 | 16.0% | 164 |
| 0.4 | 0.6 | 0.432 | 0.36 | 0.1866 | 2.45 | 17.3% | 254 |
| $P_{\text{dual}} = P_{\text{single}} * P_{\text{single}} * 1.4$ | | | | | | | |
| 0.7 | 0.3 | 0.126 | 0.09 | 0.0159 | 6.13 | 7.4% | 202 |
| 0.6 | 0.4 | 0.192 | 0.16 | 0.0502 | 3.61 | 11.0% | 221 |
| 0.5 | 0.5 | 0.35 | 0.25 | 0.1225 | 2.39 | 12.75% | 365 |
| 0.4 | 0.6 | 0.504 | 0.36 | 0.2540 | 1.65 | 10.1% | 1223 |

**Table A3.2.** Power to detect a 15% drop in sensitivity (or higher drop) over a two-day period with a nose-only swabbing LFT brand (when compared to a nose-throat swabbing LFT from a different manufacturer with equivalent device sensitivity) with 5% significance level.

| LFT Sensitivity (nose-throat) | LFT sensitivity (nose-throat) over two days | LFT sensitivity (nose-only) over two days | Number of positive PCR cases | Power |
| --- | --- | --- | --- | --- |
| 0.7 | 0.91 | 0.77 (=0.91*0.85) | 200 | 96% |
| 0.6 | 0.84 | 0.714 | 200 | 85% |
| 0.5 | 0.75 | 0.638 | 200 | 68% |
| 0.4 | 0.64 | 0.555 | 200 | 50% |
| 0.7 | 0.91 | 0.77 | 250 | 98% |
| 0.6 | 0.84 | 0.714 | 250 | 92% |
| 0.5 | 0.75 | 0.638 | 250 | 77% |
| 0.4 | 0.64 | 0.555 | 250 | 58% |
| 0.7 | 0.91 | 0.77 | 300 | 99% |
| 0.6 | 0.84 | 0.714 | 300 | 96% |
| 0.5 | 0.75 | 0.638 | 300 | 85% |
| 0.4 | 0.64 | 0.555 | 300 | 66% |

## Appendix 4: Viral Culture and Bioinformatics Analysis

Swabs were collected by the study team and transported directly to a University of Liverpool Containment Level 3 (CL3) facility. The universal transport media (UTM) was split into cryovials with μL aliquots. aliquot used for viral culture and the remainder frozen at −80°C. Calu3 cells, cultured at 10^5 cells/well in 24 well plates, were inoculated for viral culture with 100μl UTM after filtration using a 0.2μm filter and centrifugation at 12000 x g for 4mins to remove bacterial contaminants. The filtered sample was diluted 1:1 with Dulbecco’s Modified Eagle Medium (DMEM) +2% Fetal Bovine Serum (FBS) + Plasmocin (clarithromycin)/Gentamicin/Amphotericin B. A mock control of medium only was on each plate. The remaining filtered sample aliquot was frozen down.

After incubation for 30 minutes at 37°C, in 5% CO2, 500μL E /2% FBS with Plasmocin/Gentamicin/Amphotericin was added to each well and plates returned to the incubator at 37°C, 5% CO2 for 3 days.

After 3 days plates were checked for cytopathic effect (CPE). If CPE was a parent 250μL of supernatant was removed from the late and added to 750μLTrizol-LS for RNA extraction. Any remaining supernatant was stored at −80°C. If no CPE was visible after 3 days, 250μL of the supernatant was taken from the well and added to a fresh well of Calu3 cells for a 2^nd^ passage. The plate was placed back into the incubator at 37°C, 5% Co2 for a further 3 days. If after days there was still no CPE visible, 250μL was taken from each well and added to 750μL Trizol-LS for RNA extraction. Any remaining supernatant was stored at −80°C.

For RNA Extraction, Thermofisher’s hasemaker™ tubes and TRizol reagent was used, following the manufacturer’s instructions. After precipitation and drying, the RNA was finally resuspended in 50μL of RNase-free water and incubated in a heat block at 60°C for 15 minutes. RNA was used for amplicon sequencing by MinION, on an Oxford Nanopore GridION device using the ARTIC V4.1 primer scheme and ligation kits (SQK-LSK109). [A4i] The workflow is summarised in Figure A4.1.

**Figure A4.1:**
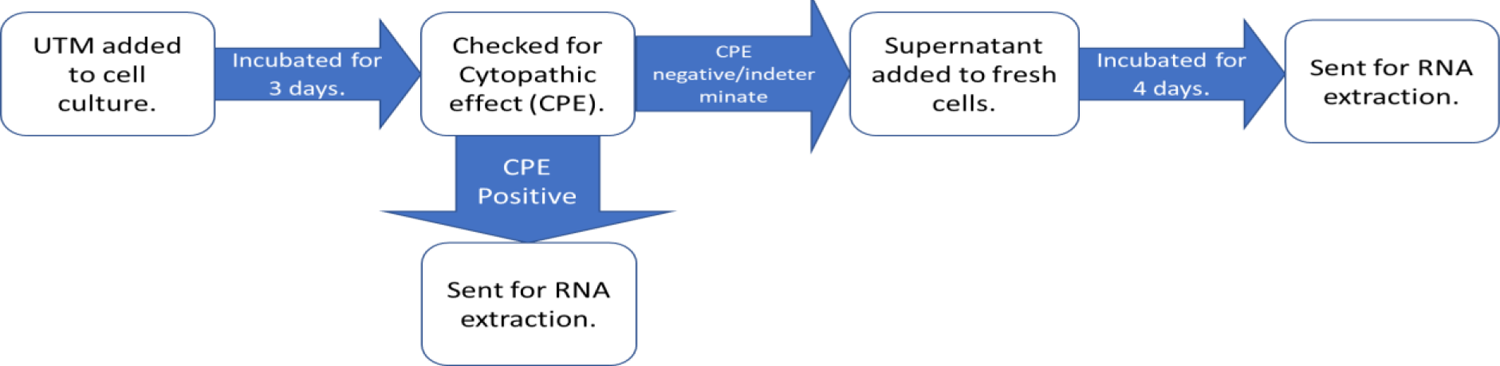
Viral culture laboratory process

A total of 41 Patients had swabs taken for laboratory analysis. 31 came from patients still testing LFT positive at Day 5-7. 10 came from patients testing LFT negative at Day 5-7.

Cultures were considered positive if CPE was present. Because CPE is not always present in viral cultures, especially with the omicron variant, culture supernatants had RNA isolated and sequenced culture supernatants had RNA isolated and sequenced by MinION. If cultures were indeterminate for CPE, or if N gene sub genomic RNA (sgRNA) was identified using a published method.[A4ii] In one case, both indeterminate CPE and N sgRNA were identified. This sample had multiple other sgRNAs detected and was considered positive. The remainder of the samples had either indeterminate CPE or N sgRNA but not both, lower levels of N sgRNA, and fewer sgRNAs detected. In these cases we were unable to prove the presence of SARS-CoV-2, these have therefore been considered indeterminate by either CPE or N sgRNA. Table A4 shows a summary of these results.

**Table A4.1:** Classification of culture result by LFT status. CPE = cytopathic effect. Bioinformatics Analysis.

| Culture result | LFT result | Laboratory result | N |
| --- | --- | --- | --- |
| Positive | Positive | CPE positive | 5 |
|  |  | CPE indeterminate, N sgRNA positive, 4 sgRNAs | 1 |
| Indeterminate | Positive | Indeterminate CPE | 9 |
| | | N sgRNA positive (low level, $\leq 3$ sgRNAs) | 2 |
|  | Negative | Indeterminate CPE | 2 |
| | | N sgRNA positive (low level, $\leq 3$ sgRNAs) | 2 |
| Negative | Positive | All negative | 14 |
|  | Negative | All negative | 6 |

## Bioinformatics Analysis

Fastq reads were analysed using the ARTIC[A4i] bioinformatic pipeline and lineages were called with Pangolin.[A4iii] LeTRS[A4ii] was used to assess the presence of sgmRNA, indicative of active viral transcription. Sequencing reads were mapped onto a known SARS-CoV-2 genome (GenBank sequence accession: NC_045512.2) using Minimap2 v 2.24-r1122 with parameters “-a -x map-ont”.[A4iv] The sorted output was used to count the number of reads ma ed on the virus genome using tools v. with “flagstat” o tion.[A4v] The “genomecov” function in edtools v2.29.2 was used to calculate the coverage of reads mapped on the virus.[A4vi] LeTRS v2.2.1 was applied to analyse sub-genomes in the sequenced samples with the options set to “-pool 0 -Rtch cDNA -mode nano ore”.[A4ii]

[A4i] SARS-CoV-2 V4.1 update for Omicron variant - Laboratory. ARTIC Real-Time Genomic Surveillance. Available at https://community.artic.network/t/sars-cov 2-v4-1-update-for-omicron-variant/342. Accessed October 10, 2023, 2021.
[A4ii] Dong X, Penrice-Randal R, Goldswain H, Prince T, Randle N, Donovan-Banfield I, et al. Analysis of SARS-CoV-2 known and novel sub-genomic mRNAs in cell culture, animal model, and clinical samples using LeTRS, a bioinformatic tool to identify unique sequence identifiers. *GigaScience* 2022;11:giac045. Doi: 10.1093/gigascience/giac045.
[A4iii] O’Toole Á, Pybus OG, Abram ME, Kelly EJ, Rambaut A. Pango lineage designation and assignment using SARS-CoV-2 spike gene nucleotide sequences. *BMC Genomics* 2022;23(1):121. Doi: 10.1186/s12864-022-08358-2.
[A4iv] Li H. Minimap2: pairwise alignment for nucleotide sequences. *Bioinformatics* 2018;34(18):3094–100. Doi: 10.1093/bioinformatics/bty191.
[A4v] Li H, Handsaker B, Wysoker A, Fennell T, Ruan J, Homer N, et al. The Sequence Alignment/Map format and SAMtools. *Bioinformatics* 2009;25(16):2078–9. Doi: 10.1093/bioinformatics/btp352.
[A4vi] Quinlan AR, Hall IM. BEDTools: a flexible suite of utilities for comparing genomic features. *Bioinformatics* 2010;26(6):841–2. Doi: 10.1093/bioinformatics/btq033.

