## Supplementary material for "Can self-testing be enhanced to hasten safe return of healthcare workers in pandemics? Random order, open label trial using two manufacturers’ SARS-CoV-2 lateral flow devices concurrently": Graphical abstract

- December 2021: Omicron → **Dangerous** levels of **key worker absence** across the UK
- Urgent response: **Use** SARS-CoV-2 **lateral flow** self-test kits **smarter** to **quicken safe return** to work

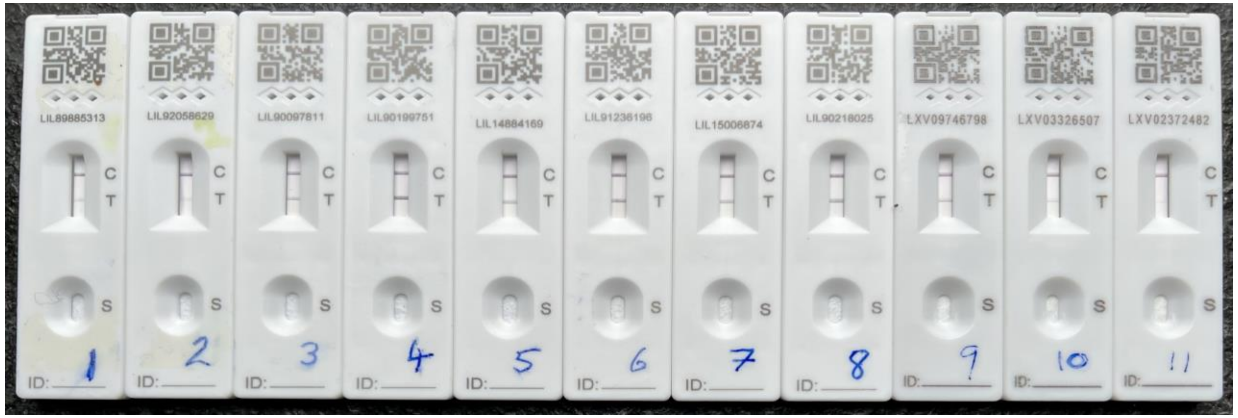


- Does dual kit testing **detect more** ongoing infection than single kit**?**
- Is dual kit testing **acceptable** to users**?**

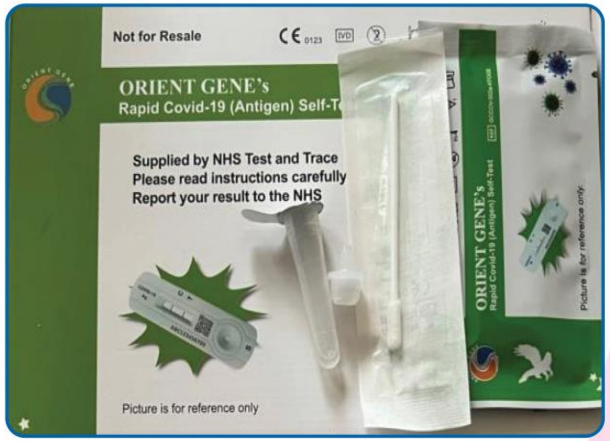


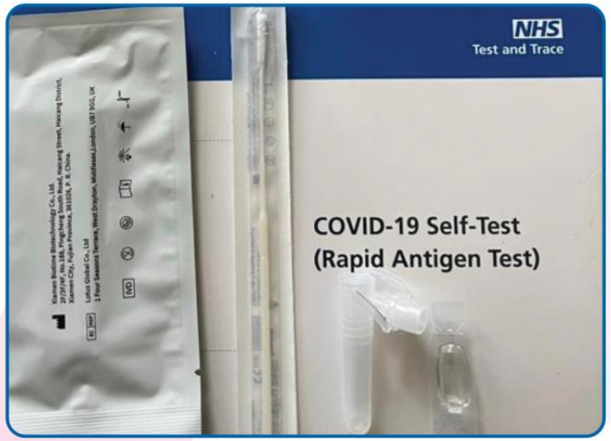


### Random order, open label dual lateral flow test trial in NHS hospital staff February-June 2022, Liverpool, UK

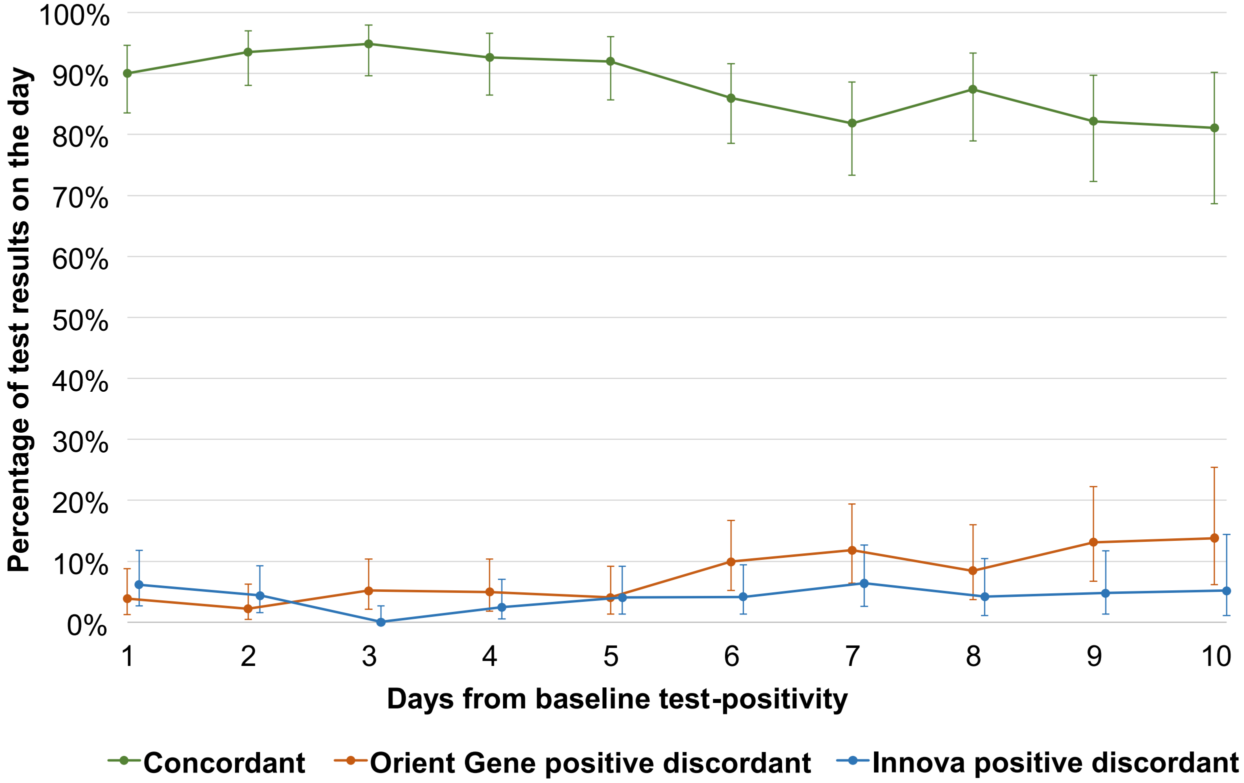


- **Improved detection**
- 91% found it **easy**; 66% **preferred** dual vs single test

Nested study of **viral culture** from swabs taken Days 5-7

| Positive (19%) | Intermediate (36%) | Negative (45%) |
| --- | --- | --- |

https://www.medrxiv.org/content/10.1101/2024.04.04.24305332v1
